# Simulating and Forecasting the Cumulative Confirmed Cases of SARS-CoV-2 in China by Boltzmann Function-based Regression Analyses

**DOI:** 10.1101/2020.02.16.20023564

**Authors:** Xinmiao Fu, Qi Ying, Tieyong Zeng, Tao Long, Yan Wang

## Abstract

An ongoing outbreak of atypical pneumonia caused by the 2019 novel coronavirus (SARS-CoV-2) is hitting Wuhan City and has spread to other provinces/cities of China and overseas. It very urgent to forecast the future course of the outbreak. Here, we provide an estimate of the potential total number of confirmed cases in mainland China by applying Boltzmann-function based regression analyses. We found that the cumulative number of confirmed cases from Jan 21 to Feb 14, 2020 for mainland China, Hubei Province, Wuhan City and other provinces were all well fitted with the Boltzmann function (*R*^*2*^ being close to 0.999). The potential total number of confirmed cases in the above geographic regions were estimated at 95% confidence interval (CI) as 79589 (71576, 93855), 64817 (58223, 77895), 46562 (40812, 57678) and 13956 (12748, 16092), respectively. Notably, our results suggest that the number of daily new confirmed cases of SARS-CoV-2 in mainland China (including Hubei Province) will become minimal between Feb 28 and Mar 10, 2020, with 95% CI. In addition, we found that the data of cumulative confirmed cases of 2003 SARS-CoV in China and Worldwide were also well fitted to the Boltzmann function. To our knowledge this is the first study revealing that the Boltzmann function is suitable to simulate epidemics. The estimated potential total number of confirmed cases and key dates for the SARS-CoV-2 outbreak may provide certain guidance for governments, organizations and citizens to optimize preparedness and response efforts.

## Introduction

An outbreak of atypical pneumonia caused by the zoonotic 2019 novel coronavirus (SARS-CoV-2) is on-going in China [1]. As of Feb 12, 2020 (24:00, GMT+8), there have been 59901 confirmed patients and 1368 deaths from SARS-CoV-2 infection, in China, and the most affected city, Wuhan, and related regions in Hubei province of China have reported 48206 confirmed patients and 1310 deaths. Cases infected in Wuhan were also detected in other provinces of China as well as in many foreign countries or regions including Japan, the Republic of Korea, Canada, USA, and European countries [2-4]. This SARS-CoV-2 outbreak was declared as a public health emergency of international concern by the World Health Organization (WHO) on Jan 30 [5].

Much research progress has been made in dissecting the evolution and origin of SARS-CoV-2 [6-8], as well as characterizing its clinical features [9-15] and epidemics [16-19] in the past one and half months. These efforts would significantly guide us to contain the SARS-CoV-2 epidemic. While the outbreak is on-going, people raise grave concerns about the future trajectory of the outbreak, especially given that the working and schooling time has been already dramatically postponed after the Chinese Lunar New Year holiday was over (scheduled on Jan 31). It is highly demanding to estimate the potential total number of confirmed cases, both nationally and locally.

Here we present Boltzmann function-based regression analyses on the data of confirmed cases of SARS-CoV-2 in China. Results indicate that the daily reported cumulative number of confirmed cases of SARS-CoV-2 regarding every defined region in China (including Hubei Province, Wuhan city, other top-6 most affected provinces and top-4 major cities) could be fitted well with Boltzmann function. Subsequent forecasting the trend of cumulative confirmed cases in each region can be made, which may be helpful for governments, organizations and citizens to optimize their preparedness and response efforts.

## Methods

### Sources of data

We collected the daily cumulative number of confirmed cases (from Jan 21, 2020 to Feb 14, 2020) infected by SARS-CoV-2 from official websites of the National Health Commission of China and of health commissions of provinces, municipalities and major cities. Overseas data were not included in our simulation due to the small number of confirmed cases. The cumulative number of confirmed cases of 2003 SARS in China and worldwide were obtained from the official website of WHO.

### Data fitting with Boltzmann function and estimation of critical dates

Data were organized in Microsoft Excel and then incorporated into Microcal Origin software (note: 2021 Jan 21 was set as day 1 and so on). The Boltzmann function was applied to data simulation for each set of data regarding different geographic regions (e.g.., China, Hubei Province and so on) and parameters of each function were obtained, with the potential total number of confirmed cases being directly given by parameter A_2_. Estimation of critical dates was performed by predicting the cumulative number of confirmed cases in the coming days post Feb 14, 2020, and the key dates were provisionally set when the number of daily new confirmed cases is lower than 0.1% of the potential total number. The Boltzmann function for simulation is expressed as follows:

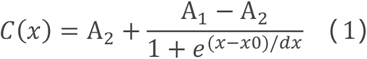

where *C(x)* is the cumulative number of confirmed cases at day *x*; A_1_, A_2_, *x0*, and *dx* are constants. In particular, A_2_ represents the estimated potential total number of confirmed cases of SARS-CoV-2. Details of derivation of the Boltzmann function for epidemic analysis are described in the supporting information file.

### Estimation of uncertainty in the non-linear regression

A Monte Carlo technique is applied to assess the uncertainty in the estimated total number of confirmed cases due to the uncertainty in the reported number cases. 1000 non-linear regressions were performed with the same time series data but each data point in the time series was perturbed by multiplying with a random scaling factor that represents the relative uncertainty. We assumed that the relative uncertainty follows a single-sided normal distribution with a mean of 1.0 and a standard deviation of 10%. This implies that all reported cases are positive but there is a tendency to miss-reporting some positive cases so that the reported numbers represent a lower limit. The resulting mean and 95% confidence interval (CI) were presented.

## Results

### Data collection and Assessment

In light of daily reported cases of SARS-CoV-2 since Jan 21, 2020, we decided to collect data for analysis on the cumulative number of confirmed cases (initially from Jan 21 to Feb 10, 2020) in several typical regions of China, including the center of the outbreak (i.e. Wuhan City and Hubei Province), other top provinces ranking in the number of cases (i.e., Guangdong, Zhejiang, Henan, Hunan, Anhui, and Jiang Provinces) and top-4 major cities in China (i.e., Beijing, Shanghai, Guangzhou, Shenzhen).

During data analysis on Feb 13, 2020, the number of new confirmed cases on Feb 12 in Hubei Province and Wuhan City suddenly increased by 14840 and 13436, respectively, of which 13332 and 12364 are those confirmed by clinical features (note: all the number of confirmed cases released by Feb 12 were counted according to the result of viral nucleic acid detection rather than by referring to clinical features). Afterward, new confirmed cases determined by clinical criteria in Hubei Province on Feb 13 and 4 are still up to 2052 and 1138. In view of these, we arbitrarily distributed these suddenly added cases to the reported cumulative number of confirmed cases from Jan 21 to Feb 14 for Hubei Province by a fixed factor (refer to **Table S1**), assuming that these newly added cases were linearly accumulative in those days. It is the same forth with the data for Wuhan City.

### Fitting data on the confirmed cases of SARS-CoV-2 to Boltzmann function and estimating the potential total number of confirmed cases

Fitting analyses using Boltzmann function indicate that all sets of data were well fitted with the function (all *R*^*2*^ values being close to 0.999; **Figs. 1, 2** and **S1**). Parameter A_2_ in the Boltzmann function directly represents the potential total number of confirmed cases (refer to **equation 1**). As summarized in **Table 1**, the potential total number of confirmed cases for mainland China, Hubei Province, Wuhan City, and other provinces were estimated as 72800±600, 59300±600, 42100±700 and 12800±100; respectively (also refer to **Fig. 1**); those for the six mostly influenced provinces (Guangdong, Zhejiang, Henan, Hunan, Anhui and Jiangxi) were 1300±10, 1170±10, 1260±10, 1050±10, 1020±10 and 940±10, respectively (also refer to **Fig. 2**); those for the top-4 major cities (Beijing, Shanghai, Guangzhou and Shenzhen) were 394±4, 328±3, 337±3 and 397±4, respectively (also refer to **Fig. S1**).

**Table 1.**
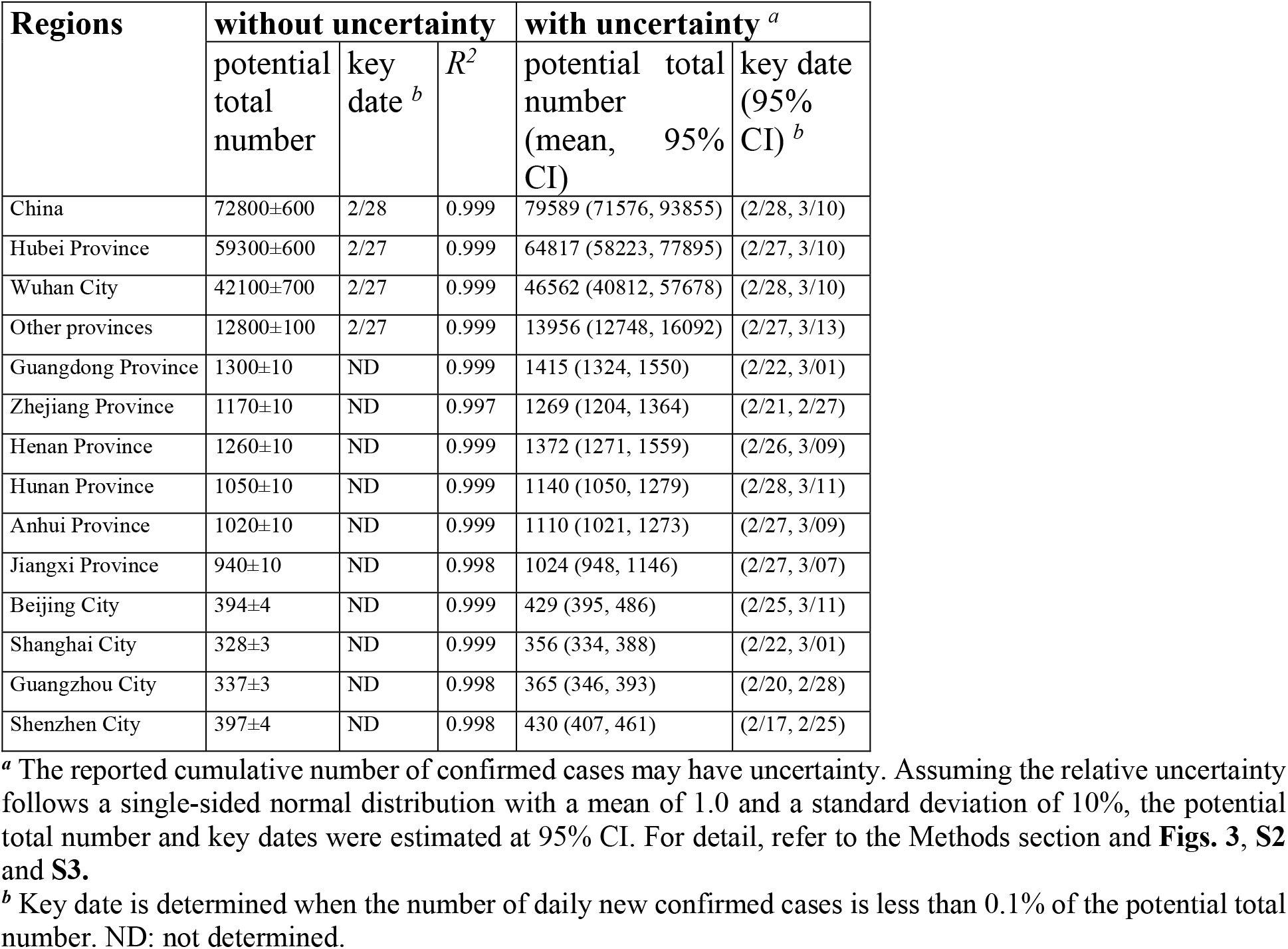
Regression analysis results of confirmed cases of SARS-CoV-2 in China.

**Figure 1.**
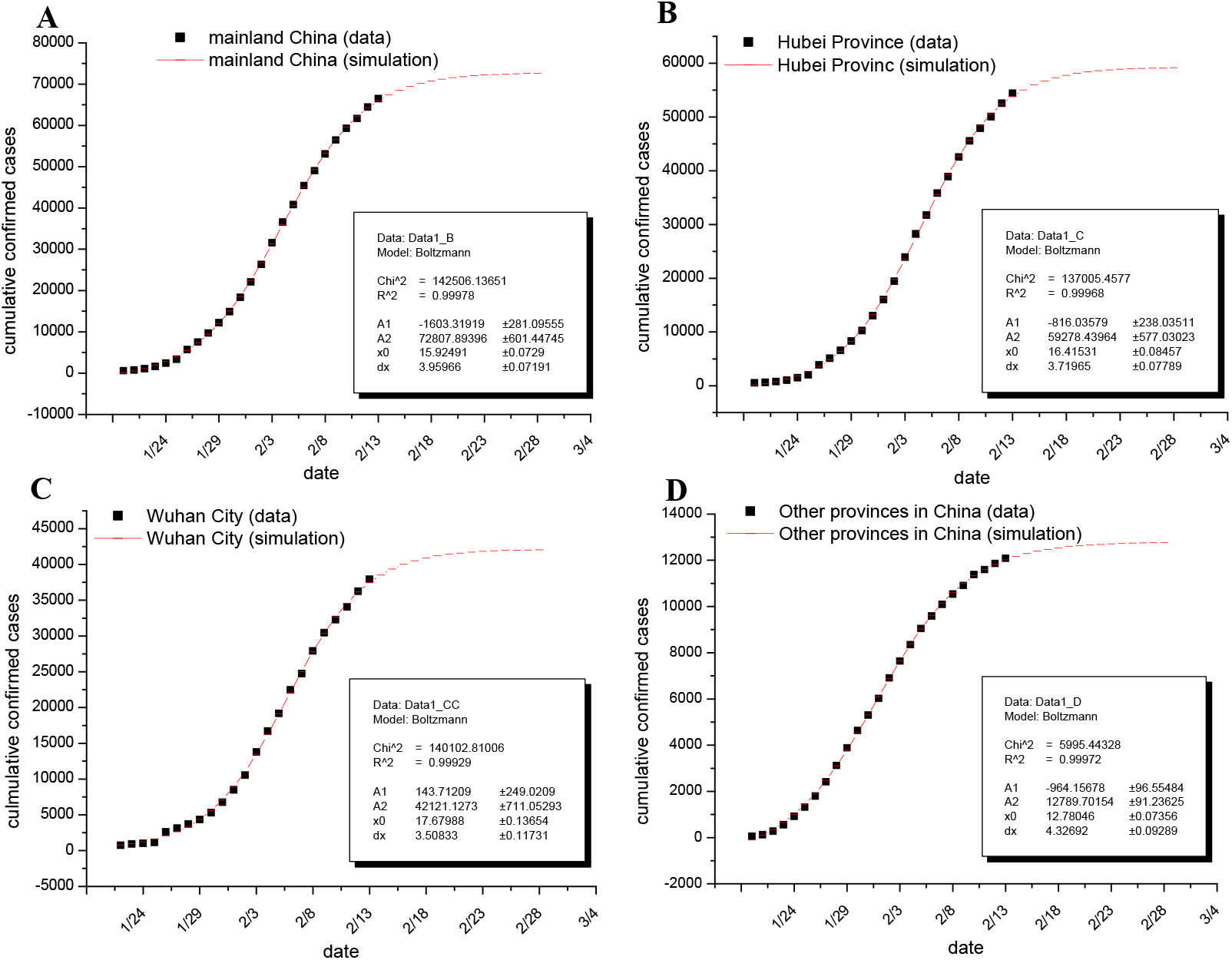
Fitting the cumulative number of confirmed cases from mainland China to Boltzmann function. Cumulative number of confirmed cases of SARS-CoV-2 as of Feb 14, 2020, in mainland China (panel A), in Hubei Province (panel B), in Wuhan City (panel C) and in other provinces (panel D) are shown as black squares, and the simulation results from Boltzmann function are plotted as red short lines and parameters of each established function are shown in inserts. Note: the reported cumulative number of confirmed cases of Hubei Province and Wuhan City were re-adjusted for data fitting due to the suddenly added cased determined by clinical features (for detail, refer to the Results section and **Table S1**).

**Figure 2.**
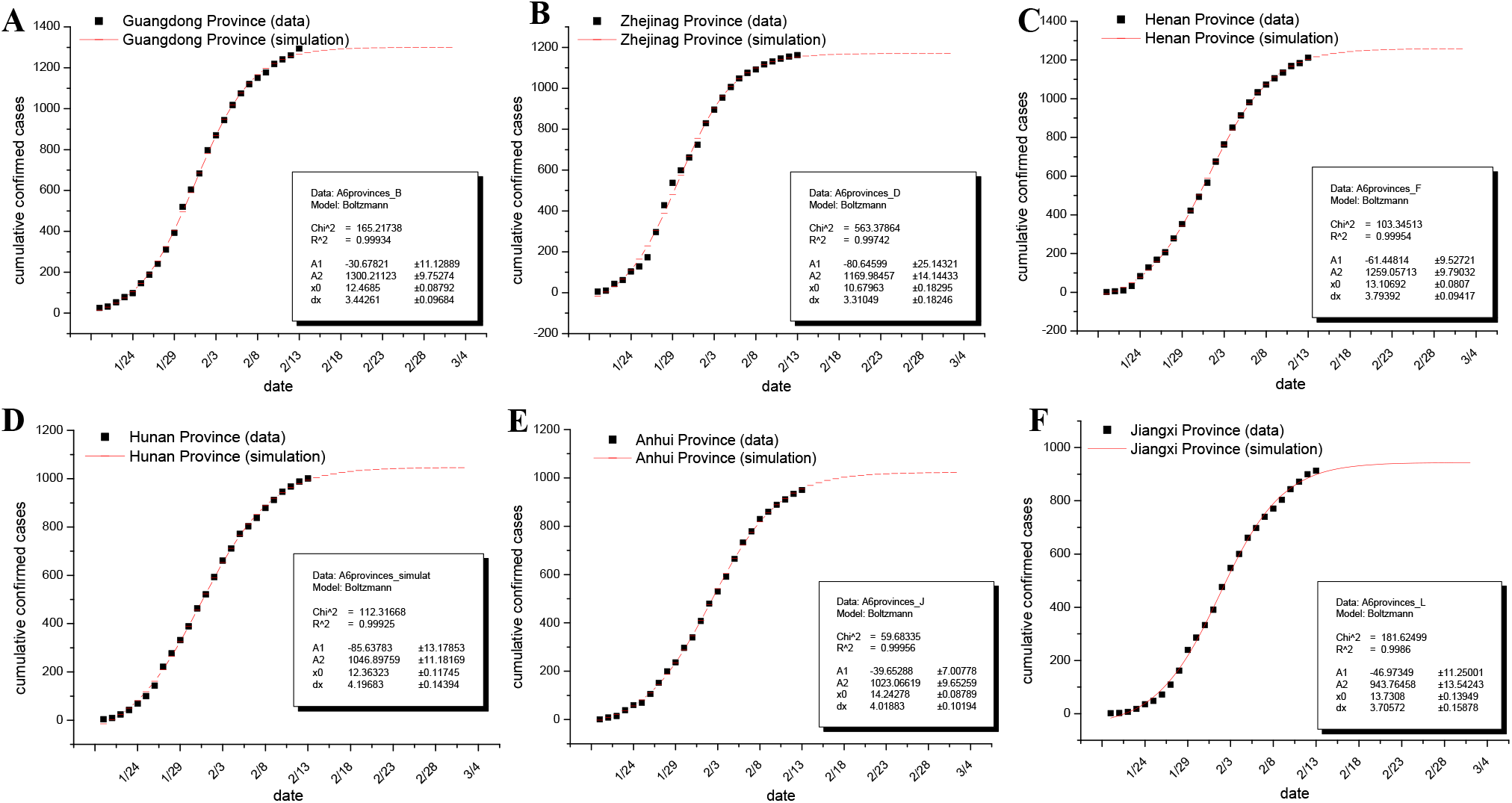
Fitting the cumulative number of confirmed cases from the six most affected provinces to Boltzmann function. Cumulative number of confirmed cases of SARS-CoV-2 as of Feb 14, 2020, in provinces Guangdong (panel A), in Zhejiang (panel B), in Henan (panel C), in Hunan (panel D), in Anhui (panel E) and in Jiangxi (panel F) are shown as black squares, and the simulation results from Boltzmann function are plotted as red short lines and parameters of each established function are shown in inserts.

In addition, we estimated the key date, on which the number of daily new confirmed cases is lower than 0.1% of the potential total number of confirmed cases as defined by us subjectively. As summarized in **Table 1**, the key dates for mainland China, Hubei Province, Wuhan City and other provinces are Feb28 or Feb 27. It appears that it will take approximately two weeks for mainland China to reach this state such that the number of daily new confirmed cases of SARS-CoV-2 post the critical date is below 70.

### Estimation of uncertainty in the non-linear regression

The above analyses were performed assuming that the released data on the confirmed cases are precise. However, there is a tendency to miss-report some positive cases such that the reported numbers represent a lower limit. One typical example indicating this uncertainty is the sudden increase of more than 13 000 new confirmed cases in Hubei province on Feb 12 after clinical features were officially accepted as a standard for infection confirmation. Another uncertainty might result from insufficient kits for viral nucleic acid detection at the early stage of the outbreak. We thus examined the effects of the uncertainty of the released data on the estimation of the potential total number of confirmed cases using a Monte Carlo method (for detail, refer to the Methods section). For simplicity, we assumed that the relative uncertainty of the reported data follows a single-sided normal distribution with a mean of 1.0 and a standard deviation of 10%.

Under the above conditions, the potential total numbers of confirmed cases of SARS-CoV-2 for different regions were estimated (**Figs. 3, S2** and **S3**) and summarized in Table 1. The potential total numbers for China, Hubei Province, Wuhan City and other provinces were 79589 (95% CI 71576, 93855), 64817 (58223, 77895), 46562 (40812, 57678) and 13956 (12748, 16092), respectively (also refer to **Fig. 3**); those for the six most affected provinces (Guangdong, Zhejiang, Henan, Hunan, Anhui and Jiangxi) were 1415 (1324, 1550), 1269 (1204, 1364), 1372 (1271, 1559), 1140 (1050, 1279), 1110 (1021, 1273) and 1024 (948, 1146), respectively (also refer to **Fig. S2**); those for top-4 major cities (Beijing, Shanghai, Guangzhou and Shenzhen) were 429 (395, 486), 356 (334, 388), 365 (346, 393) and 430 (407, 461), respectively (also refer to **Fig. S3**).

**Figure 3.**
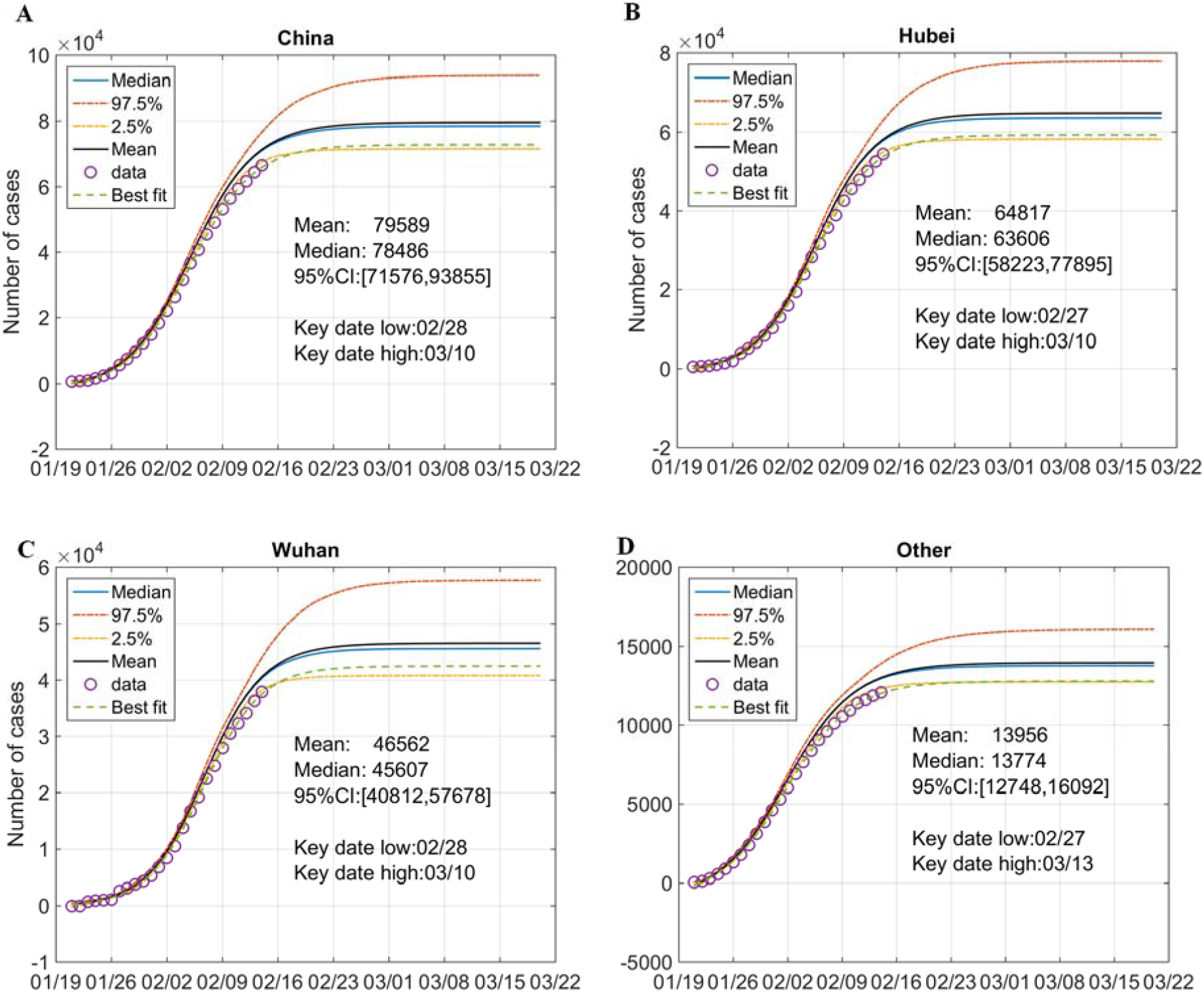
Analysis of the uncertainty of confirmed cases for mainland China, Hubei Province, Wuhan City and other provinces. Data of mainland China (panel A), of Hubei Province (panel B), of Wuhan City (panel C) and of other provinces (panel D) were fitted to Boltzmann function assuming that the relative uncertainty of the data follows a single-sided normal distribution with a mean of 1.0 and a standard deviation of 10%. Original data are shown as circles; simulated results are presented as colored lines as indicated. Inserts show key statistics. The key date is defined as the date when the number of daily new confirmed cases is less than 0.1% of the potential total number. The low and high key dates were determined by the simulated curve of confidence interval (CI) at 2.5% and 97.5%, respectively.

Such uncertainty analysis also allowed us to estimate the key dates at 95% CI. As summarized in **Table 1**, the key dates for mainland China, Hubei Province, Wuhan City, and other provinces would fall in (2/28, 3/10), (2/27, 3/10), (2/28, 3/10) and (2/27, 3/13), respectively (also refer to **Fig. 3**); those for the six provinces (Guangdong, Zhejiang, Henan, Hunan, Anhui and Jiangxi) were within (2/22, 3/01), (2/21, 2/27), (2/26, 3/09), (2/28, 3/11), (2/27, 3/09) and (2/27, 3/07), respectively (also refer to **Fig. S2**); those for top-4 major cities (Beijing, Shanghai, Guangzhou and Shenzhen) were within (2/25, 3/11), (2/22, 3/01), (2/20, 2/28) and (2/17, 2/25), respectively (also refer to **Fig. S3**).

### Data on the confirmed cases of 2013 SARS-CoV were well fitted to Boltzmann function

The ongoing SARS-CoV-2 outbreak has undoubtedly caused the memories of the SARS-CoV outbreak in 2003. Prompted by the above observation that the data of the SARS-CoV-2 outbreak so far were well fitted to Boltzmann function, we thus applied this function to fit the epidemic of 2003 SARS-CoV in China and worldwide. Results in **Fig. 4** show that the cumulative numbers of confirmed cases of 2003 SARS-CoV both in China and worldwide were fitted well with the Boltzmann distribution function, with *R*^*2*^ being 0.999 and 0.998, respectively.

**Figure 4.**
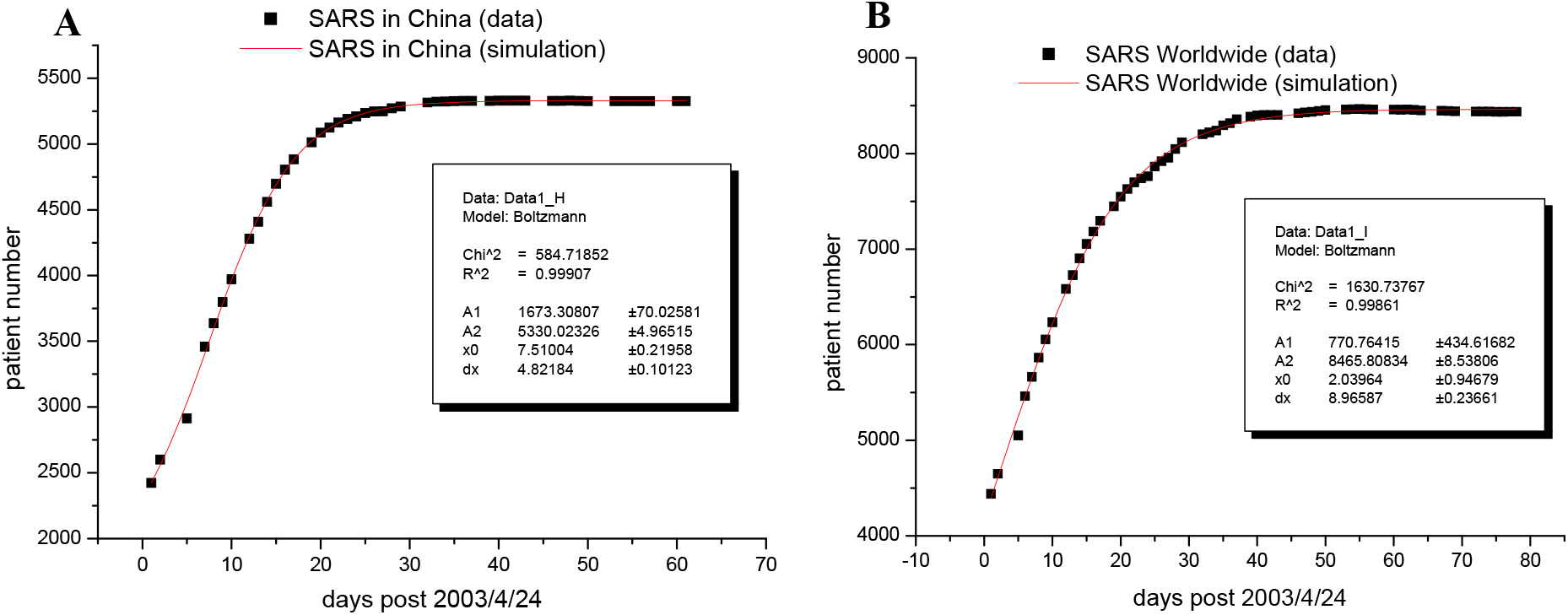
Fitting the cumulative number of confirmed cases for 2003 SARS in China and worldwide to Boltzmann function. The cumulative number of confirmed cases of 2003 SARS in China (panel A) and worldwide (panel B) are shown as black squares, and the simulation results from Boltzmann function are plotted as red short lines and parameters of each established function are shown in inserts.

## Discussion and conclusion

It is of significance to estimate the spread of the SARS-CoV-2 outbreak originating in Wuhan, China. In particular, a precise estimation of the potential total number of infected cases and/or confirmed cases is critical for optimal preparedness and implementation of infection control practices made by governments nationally and locally. An earlier study published on 17 January 2020 [18] suggested that a total of 1,723 cases of SARS-CoV-2 in Wuhan City (95% CI: 427-4,471) had onset of symptoms by 12th January 2020, which was calculated based on the internationally reported cases. Later, Wu et al [19] applied a susceptible-exposed-infectious-recovered (SEIR) metapopulation model to simulate the epidemics of the SARS-CoV-2 outbreak across all major cities in China on the basis of the number of cases exported from Wuhan to cities outside mainland China, and they estimated that 75 815 individuals (95% CI 37 304–130 330) had been infected in Wuhan as of Jan 25, 2020. A recent study based on the Susceptible-Infected-Recovered-Dead (SIRD) model revealed that the cumulative number of infected cases will surpass 68,000 (as a lower bound) and could reach 140,000 (with an upper bound of 290,000) by February 29 [17]. Notably, Huang and Qiao proposed a simple and data driven model based on the physics of natural growth algorithm and estimated the transmission rate of the SARS-CoV-2 outbreak [16]. The results of these modeling studies have provided important guidance for governments and health agencies to intensify preparedness and response efforts.

Here we applied Boltzmann function to analyze the reported confirmed cases in mainland China, focusing Hubei Province and Wuhan City, the center of the outbreak, as well as those most affected provinces and economic centers. Strikingly, all data sets were well fitted to the Boltzmann function (**Figs. 1, 2** and **S1**). More importantly, the data of 2003 SARS-CoV in China and worldwide were also well fitted to the function (**Fig. 4**). These results, in conjunction with that Boltzmann function can be inferred from a few assumptions (for detail, refer to the Methods section of the support information file), suggest that Boltzmann function is suitable for analyzing the epidemics of coronaviruses like SARS-CoV and SARS-CoV-2. One advantage of this model is that parameter A_2_ directly gives an estimate of the potential total numbers of confirmed cases. In addition, unlike traditional epidemiological models that require much more detailed data for analysis [14, 19], Boltzmann function-based regression analysis only needs the cumulative number of confirmed cases, somehow as simple as the model proposed by Huang and Qiao [16].

Furthermore, the established Boltzmann functions allow us to forecast the future course of the epidemics of SARS-CoV-2 in different regions in the coming weeks. The potential total numbers of confirmed cases in mainland China, in Hubei Province, in Wuhan City (the center of the outbreak) and in other most affected provinces and top4 major cities, as summarized in **Table 1**, may provide valuable guidance for Chinese central and local governments to contain this outbreak at current critical stage. In particular, the numbers in mainland China and Hubei Province were estimated as 79589 (95% CI 71576, 93855) and 64817 (95% CI 58223, 77895), respectively, indicating that the SARS-CoV-2 outbreak in China might not be as bad as thought. Notably, our results also suggest that the number of daily new confirmed cases will become minimal between Feb 28 and Mar 10 in mainland China (including Hubei Province) at 95% CI (**Fig. 3A**). This trend, if occur as predicted, may help citizens in China to release stress and anxiety, as there have been many provinces and/cities in China that have suspended public transportation systems and even implemented house quarantines for all urban households [20]. In further support of these estimates by the Boltzmann function, the newly released cumulative number of confirmed cases in all the above geographic regions on Feb 15 and Feb 16 are very close to the predicted ones (refer to **Table S2**). Consistently, parameters of the established Boltzmann functions by regression analyses of the data from Jan 21 to Feb 16, 2020 (as presented in **Fig. S4**) are highly similar to those made by the data from Feb 21 to Feb 14, 2020.

Nevertheless, our estimates based on the established Boltzmann functions are not absolutely guaranteed, mainly because of the uncertainty of the reported data (**Figs. 3, S2** and **S3**). We estimated the potential total numbers (refer to **Table 1**) under the assumption that the relative uncertainty of the reported data follows a single-sided normal distribution with a mean of 1.0 and a standard deviation of 10%, and this deviation may be underestimated. If the real uncertainty of released data by health commissions is larger than 10%, the potential total numbers of confirmed cases would accordingly increase, and the key dates will be postponed. Another limitation is that this estimate is based on the assumption that the overall conditions are not changing. This might not be true, given that in many regions the workers have started to return for work half a month post the Spring Festival holiday (schedulely ending on Feb 31), which may increase the SARS-CoV-2 infection. In this regard, it is noted that the daily number of new confirmed cases in past a few days in several provinces and cities (e.g., Guangdong Province, **Fig. 2A**; Shanghai and Shenzhen City, **Fig. S1D**) have increased a little bit more than predicted by the model.

## Data Availability

All data are presented in the manuscript and supplementary data file.

## Acknowledgments

We thank graduate students (Boyan Lv, Zhongyan Li, Zhongyu Chen, Yu Cheng, Mengmeng Bian, Shuang Zhang, Zuqin Zhang, and Wei Yao; all from Prof. Xinmiao Fu’s research group at Fujian Normal University) for data collection from official websites of National and Health Commission of China and of provincial health commissions in China. The authors would like to acknowledge colleagues for helpful comments. This work is support by the National Natural Science Foundation of China (No. 31972918 and 31770830 to XF).

## Declarations

### Ethics approval and consent to participate

Ethical approval or individual consent was not applicable.

### Availability of data and materials

All data and materials used in this work were publicly available, and also available based on request.

### Disclaimer

The funding agencies had no role in the design and conduct of the study; collection, management, analysis, and interpretation of the data; preparation, review, or approval of the manuscript; or decision to submit the manuscript for publication.

### Author Contributions

I. Conception and design: Xinmiao Fu
II. Derivation of the Boltzmann function: Tieyong Zeng
III. Data simulation and estimation: Xinmiao Fu
IV. Uncertainty analyses: Qi Ying
V. Data collection and organization: Tao Long and Yan Wang
VI. Manuscript drafting: Xinmiao Fu

